# IMPACT OF THE COVID-19 PANDEMIC ON ADHERENCE TO CARDIOVASCULAR MEDICATIONS AMONG CHRONICALLY TREATED PATIENTS IN ALBERTA

**DOI:** 10.1101/2024.07.31.24311323

**Authors:** Finlay A. McAlister, Anamaria Savu, Luan Manh Chu, Douglas C. Dover, Padma Kaul

**Affiliations:** From: The Canadian Vigour Centre, 4-120 Katz Group Centre for Pharmacy and Health Research, University of Alberta College of Health Sciences, Faculty of Medicine & Dentistry, Edmonton, AB T6G 2E1

**Author notes:** Correspondence: Dr. F. McAlister, 5-134C Clinical Sciences Building, University of Alberta, 11350 83 Avenue, Edmonton, Alberta, Canada T6G 2G3.

## Abstract

**Background:** Some studies have suggested that the COVID-19 pandemic negatively impacted patient adherence with chronic therapies. We designed this study to explore whether cardiovascular drug adherence patterns changed in chronically treated patients during the COVID-19 pandemic.

**Methods:** Retrospective cohort study examining drug dispensation data for all Alberta residents older than 18 years who were chronic users of at least one cardiovascular drug class, defined as receiving at least one prescription per annum for any agent in that drug class from March 15, 2017 to March 14, 2023 and 2 or more prescriptions within 365 days during either the pre-pandemic phase (March 15, 2018 to March 14, 2020) or the pandemic phase (March 15, 2020 to March 14, 2022). We calculated the proportion of days covered (PDC) for each drug class per patient and used generalized estimating equation logistic regression to estimate the effect of time period (pandemic versus pre-pandemic) on achievement of good adherence (PDC>0.8) after adjusting for age, sex, socioeconomic status, and comorbidities.

**Results:** We evaluated 548,601 chronic users of at least one cardiovascular drug class between March 15, 2018 and March 14, 2022. The most frequently dispensed cardiovascular drug classes were ACEi/ARB (67.6%), statins (53.8%), beta-blockers (21.0%), and calcium channel blockers (20.7%); the most frequent diagnoses were hypertension (77.2%), diabetes mellitus (30.6%), and ischemic heart disease (19.6%), although 55.4% of the patients had Charlson Comorbidity Index scores of 0. The mean PDC for cardiovascular drug use in our cohort was 85.7% (median 93%) pre-pandemic and 87.0% (median 94%) during the pandemic. The proportion exhibiting good adherence (PDC>0.8) increased from 72.8% pre-pandemic to 75.4% during the pandemic (p<0.001). During the pandemic, users of cardiovascular drugs were more likely to exhibit good adherence (PDC>0.8) than they were in the pre-pandemic period: aOR ranged from 1.05 (95%CI 1.00-1.11) for mineralocorticoid receptor antagonists to 1.16 (1.15-1.17) for statins.

**Conclusion:** Chronic users of cardiovascular drugs exhibited higher adherence measures during the COVID-19 pandemic than prior to the pandemic.

---

It has long been recognized that adherence with proven efficacious therapies is associated with better patient outcomes and lower health care costs.[1–4] There are multiple reasons to hypothesize that the COVID-19 pandemic may have adversely affected patient adherence to long-term therapies: negative press and social media reports about some cardiovascular drugs increasing risk of COVID-19 (particularly ACEi and ARB), patient fear of going to pharmacies while COVID-19 was circulating in the community, many pharmacies reduced dispensation quantities to a maximum of 30 days from the usual 90 days to preserve the supply chain, and documented increases in drug shortages due to global supply disruptions. However, there is little empiric data published thus far. Three small surveys of selected patient samples suggesting adherence may have been substantially negatively impacted during the pandemic (with as many as one fifth of survey respondents reporting difficulty obtaining their medications early during the pandemic and up to a one third drop in adherence).[5–7] While an analysis of 27,318 American adults older than 40 years receiving antihypertensive prescriptions reported a drop in adherence from 65.6% pre-pandemic to 58.9% in the first 6 months after pandemic onset,[8] that analysis only included the period with the most widespread societal lockdowns and biggest reductions in access to outpatient care. Thus, we designed this study to explore whether cardiovascular drug adherence patterns in the Canadian province of Alberta changed during the COVID-19 pandemic beyond the initial few months – this is an important question to answer given the likelihood of future pandemics and thus the need to plan for the extent of resources that should be diverted to mitigate this issue during future pandemics.

## METHODS

### Design

We conducted a retrospective cohort study examining drug dispensation data for all adult Albertans between March 15, 2017 and March 14, 2023. COVID-19 was declared a pandemic and emergency health measures were instituted in Alberta on March 15, 2020.

### Data Sources

The Alberta Pharmacy Information Network captures all drug dispensations in outpatient pharmacies in Alberta for patients of all ages. We linked the drug dispensation data with Alberta’s Health Care Insurance Plan (AHCIP) registry files (which includes basic demographics as well as home address), physician claims database, the National Ambulatory Care Reporting System (which captures all emergency department visits in Alberta and includes up to 10 diagnoses), and the Canadian Institute for Health Information (CIHI-DAD) hospitalization database (which captures up to 25 diagnoses). Linkage was achieved using each individual’s unique encrypted health identifier number and we were able to extract patient demographics, cardiovascular diagnoses, and comorbidity profiles from the linked datasets.

### Study Samples – two time periods

We compared cardiovascular drug dispensations for all Alberta residents older than 18 years in two time periods: 1) pre-pandemic: March 15, 2018 to March 14, 2020 and 2) pandemic: March 15, 2020 to March 14, 2022. Within each time period, we identified chronic users of each of the cardiovascular drug classes of interest using the same algorithm designed to avoid the pitfalls associated with defining a patient as a medication user on the basis of a single prescription or missing dispensations within the 12 months immediately before or after each time period. Thus, we defined chronic users as those receiving at least one prescription per annum for any agent in that drug class from March 15, 2017 to March 14, 2023 with 2 or more prescriptions on different days but within 365 days during either the pre-pandemic phase (March 15, 2018 to March 14, 2020) or the pandemic phase (March 15, 2020 to March 14, 2022). This algorithm was applied to each drug class separately and ensured that we were examining chronic users, not new users or discontinuing users. Lastly, we chose only those patient-drug class combinations which met the chronic users definition in both time periods to form our study sample. We excluded patients who were not registered with the Alberta Health Care Insurance Plan between March 15, 2017 and March 14, 2023, were younger than 18 at March 15, 2017, or who emigrated or died between Mar 15, 2017 and Mar 14, 2022.

### Drug classes of interest

We examined adherence with each of 6 drug classes (eTable 1): 1) angiotensin converting enzyme inhibitor or angiotensin II receptor blocker (ACEi/ARB), 2) statin, 3) calcium channel blocker (CCB), 4) beta blocker (BB), 5) thiazide and thiazide-like diuretics (including combinations with potassium sparing agents), and 6) mineralocorticoid receptor antagonist (MRA). For patients receiving multiple drug classes concomitantly, we calculated the patient’s PDC for each drug class separately.

We excluded from consideration drugs that were not on the general formulary in Alberta and/or required special authorization processes in the timeframe studied, such as angiotensin receptor antagonist neprilysin inhibitors, sodium glucose cotransporter 2 inhibitors, glucagon-like peptide-1 receptor agonists, and direct oral anticoagulants. We also excluded drugs prescribed for short timeframes (such as P_2_Y_12_ receptor antagonists after coronary interventions), drugs that could be purchased over the counter (such as aspirin), or drugs where the daily dose could vary based on test results (such as warfarin), fluid status (such as loop diuretics), or symptoms (such as nitrates).

### Definitions of adherence

We calculated adherence for each drug class using the ‘days supplied’ variable in the Alberta Pharmacy Information Network for all dispensations during each of the study time periods, recognizing that one patient could contribute data for several drug classes. Our primary analysis was the proportion of days covered (PDC) for chronic users of each drug class, calculated for each patient-drug dyad as the total number of days supplied for all dispensations of that drug class (regardless of dose) in the numerator and the number of days between March 15^th^, 2018 and March 14^th^, 2020 (731 days) and between March 15^th^, 2020 and March 14^th^, 2022 (730 days) as the denominator in the pre-pandemic and pandemic periods, respectively. We analyzed PDC as a continuous variable as well as in strata established in the literature (>80% vs. 40-80% vs. <40% for defining good vs. fair vs. poor adherence).[1,9]

### Outcomes

Our outcomes of interest were adherence at the level of each of the 6 drug classes and at the level of the patient (averaging across all drug classes dispensed for that patient), and we compared PDC and PDC strata (good, fair, or poor) in the pre-pandemic period with those in the pandemic period. Our primary outcome was the proportion of chronic users who exhibited “good adherence” (PDC>0.8) for each drug class in the 2 years before (Mar 15, 2018-Mar 14, 2020) and after (Mar 15, 2020-March 14, 2022) the declaration of the COVID-19 pandemic in Alberta.

### Comorbidities

We used ICD-9 and ICD-10-CA case definitions previously validated in Alberta [10] for any hospitalizations, any emergency department visits, and any outpatient visits in the two years prior to and including the date of the first drug dispensation for that class in that patient to identify comorbidities for each patient (eTable 1). Of note, comorbidities identified only on outpatient visits to physician offices were considered as present only if at least two such claims were identified on two different days during the study timeframe. The Charlson Comorbidity index was calculated for each patient using the weights established by Shneeweiss et al.[11]

### Statistical Analyses

For each period (pre-pandemic and pandemic), chronic users of any of the 6-drug classes of interest were pooled together and we report descriptive statistics (means and standard deviations for continuous variables, counts and percentages for categorical variables) for chronic users in both time periods. The variables included demographics (age, sex, type of residence, material deprivation index based on home address), comorbidities (Charlson Comorbidity Index, ischemic heart disease, hypertension heart failure, diabetes), PDC, and PDC strata. A 100%-stacked bar chart was used to display differences in PDC strata for chronic users of each drug class between the pre-pandemic and pandemic time periods. For patients receiving multiple drug classes concomitantly, we calculated the patient’s overall PDC as the average of their individual drug class PDC values.

To determine whether adherence changed from pre-pandemic to pandemic we used an adjusted logistic regression model for each medication class. The outcome was good adherence (PDC>0.8 versus not) and independent variables included time period (pre-pandemic, pandemic), sex (female, male), residence (urban, rural), material deprivation index (quintiles, a proxy for socioeconomic status), Charlson Comorbidity Index (0, 1-2, 3+), presence of heart failure, ischemic heart disease, hypertension, or diabetes (yes, no), and age (continuous). As each patient had a PDC record for both pre-pandemic and pandemic, we further adjusted the models for correlated data on the same user using generalized estimating equations (GEE). Thus, for each medication class the model estimated the adjusted effect of pandemic versus pre-pandemic on adherence.

A two-tailed p-value of less than 0.05 was considered significant. All analyses were conducted using SAS Version 9.4 (SAS Institute, Cary, NC) This study was approved by the University of Alberta Health Research Ethics Board (Pro00115481) with a waiver of individual patient consent as the investigators only had access to de-identified data.

## RESULTS

There were 46,308,952 dispensations for the cardiovascular medications of interest between March 2018 and March 2022. After exclusion of prescriptions dispensed to patients who were not Alberta residents, were younger than 18, did not have at least 2 dispensations within the same drug class, or who died or emigrated during the study period, we identified 835,566 users of at least one cardiovascular drug class in the 2 years prior to pandemic onset and 889,068 in the first 2 years of the pandemic (Figure 1). After excluding user-drug class combinations which were not dispensed in all years, our study cohort consisted of 548,601 patients who were chronic users of at least one cardiovascular drug class across both the pre-pandemic and pandemic periods. The most frequently dispensed cardiovascular drug classes were ACEi/ARB (67.6%), statins (53.8%), beta-blockers (21.0%), and calcium channel blockers (20.7%) – Figure 1. The most frequent diagnoses were hypertension (77.2%), diabetes mellitus (30.6%), and ischemic heart disease (19.6%), although 55.4% of the patients had Charlson Comorbidity Index scores of 0 (Table 1).

**Figure 1.**
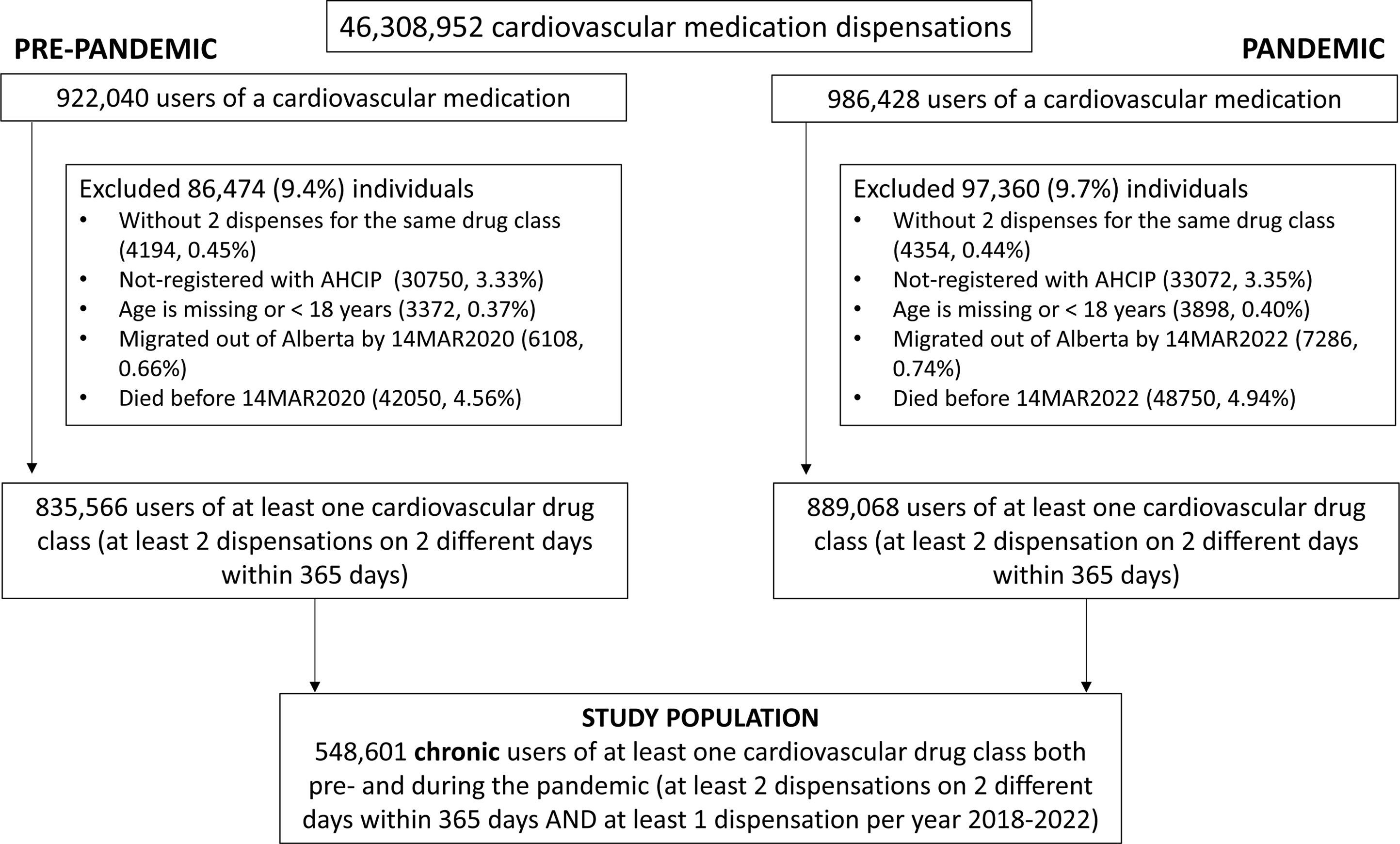
Flow Chart

**Table 1.** Characteristics of chronic users of cardiovascular drugs of interest during both study periods.

|  | Pre-pandemic<br>[Mar 15 2018 - Mar 14 2020] | Pandemic<br>[Mar 15 2020 - Mar 14 2022] |
| --- | --- | --- |
|  | 548601 | 548601 |
| Age (mean, SD) | 62.3 (12.3) | 64.3 (12.3) |
| Age 65+ | 239505 (43.7) | 274697 (50.1) |
| Female | 259035 (47.2) |  |
| Urban | 330573 (60.3) | 329559 (60.1) |
| Material Deprivation Index<br>5<sup>th</sup> quintile (most deprived) | 120045 (21.9) | 117955 (21.5) |
| 4<sup>th</sup> quintile | 118680 (21.6) | 117810 (21.5) |
| 3<sup>rd</sup> quintile | 102177 (18.6) | 102209 (18.6) |
| 2<sup>nd</sup> quintile | 92639 (16.9) | 93708 (17.1) |
| 1<sup>st</sup> quintile (most privileged) | 87317 (15.9) | 88138 (16.1) |
| Missing | 27743 (5.1) | 28781 (5.2) |
| CCI Score: |  |  |
| Mean (SD) | 0.9 (1.4) | 1.0 (1.5) |
| CCI 0 | 303707 (55.4) | 285112 (52.0) |
| CCI 1 or 2 | 187525 (34.2) | 192999 (35.2) |
| CCI 3 or more | 57369 (10.5) | 70490 (12.8) |
| Comorbidities based on all encounters in the 24 months up to time of first prescription in each time period (n, %): |  |  |
| Heart Failure | 30186 (5.5) | 36317 (6.6) |
| Ischemic Heart Disease | 107501 (19.6) | 111212 (20.3) |
| Hypertension | 423291 (77.2) | 432021 (78.7) |
| Diabetes Mellitus | 167598 (30.6) | 181426 (33.1) |
| Drug Class: |  |  |
| ACEi or ARB | 370945 (67.6) |  |
| Statin | 295421 (53.8) |  |
| CCB | 113551 (20.7) |  |
| BB | 115011 (21.0) |  |
| Thiazide | 29465 (5.4) |  |
| MRA | 9951 (1.8) |  |
| Adherence: |  |  |
| Mean PDC (SD) | 85.7 (16.7) | 87.0 (16.1) |
| Median PDC (IQR) | 93 (78, 98) | 94 (80, 99) |
| PDC < 0.4 (poor) | 11740 (2.1) | 9856 (1.8) |
| PDC 0.4 to 0.8 (fair) | 137507 (25.1) | 125229 (22.8) |
| PDC ≥ 0.8 (good) | 399354 (72.8) | 413516 (75.4) |

CCI – Charlson Comorbidity Index, ACEi – angiotensin-converter enzyme inhibitor, ARB - angiotensin II receptor blocker, BB - beta blocker, CCB - calcium channel blocker, MRA – mineralocorticoid receptor antagonist, PDC – proportion of days covered, SD – standard deviation, IQR – interquartile range, Unless otherwise specified reported values are counts (percentage)

The number of prescriptions dispensed and the number of unique users per annum increased steadily across all study years for each drug class except thiazides (which were declining even prior to the pandemic) with no evident step changes at the time of pandemic onset in mid-March 2020 (Table 2). The annual number of dispensations for each drug class varied between 5 and 6 per user across all years studied (Table 2).

**Table 2.** Number of prescription dispensations and unique users for cardiovascular drug classes by year in Alberta.

|  |  | **PRE-PANDEMIC**<br>**Study Period** |  | **PANDEMIC Study**<br>**Period** |  |  | Number of patients<br>meeting chronic user<br>definition in each year |
| --- | --- | --- | --- | --- | --- | --- | --- |
| Medication<br>class | Mar 15,<br>2017 to<br>Mar 14,<br>2018 | Mar 15,<br>2018 to<br>Mar 14,<br>2019 | Mar 15,<br>2019 to<br>Mar 14,<br>2020 | Mar 15,<br>2020 to<br>Mar 14,<br>2021 | Mar 15,<br>2021 to<br>Mar 14,<br>2022 | Mar 15,<br>2022 to<br>Mar 14,<br>2023 |  |
| ACEiARB |  |  |  |  |  |  |  |
| Total<br>Dispensations | 2992012 | 3047769 | 3213203 | 3882531 | 3423555 | 3449466 |  |
| Unique users | 536190 | 558816 | 582735 | 590503 | 611124 | 636067 | 370945 |
| STATIN |  |  |  |  |  |  |  |
| Total<br>Dispensations | 2284528 | 2317867 | 2496678 | 2982938 | 2814626 | 2933203 |  |
| Unique users | 428075 | 451866 | 476386 | 494717 | 529811 | 561220 | 295421 |
| CCB |  |  |  |  |  |  |  |
| Total<br>Dispensations | 1188230 | 1202978 | 1300223 | 1505666 | 1409162 | 1446323 |  |
| Unique users | 205941 | 217661 | 230415 | 238544 | 249025 | 262871 | 113551 |
| BB |  |  |  |  |  |  |  |
| Total<br>Dispensations | 1378103 | 1359105 | 1438667 | 1603633 | 1505069 | 1509875 |  |
| Unique users | 210597 | 217073 | 224807 | 226559 | 233724 | 240752 | 115011 |
| N | 173148 | 188209 | 200788 | 207157 | 206132 | 184785 |  |
| THIAZIDE |  |  |  |  |  |  |  |
| Total<br>Dispensations | 387826 | 365273 | 372418 | 397957 | 327010 | 309773 |  |
| Unique users | 85621 | 83890 | 87092 | 76059 | 70306 | 67523 | 29465 |
| MRA |  |  |  |  |  |  |  |
| Total<br>Dispensations | 183377 | 190287 | 215657 | 255586 | 263701 | 282799 |  |
| Unique users | 30905 | 33910 | 37764 | 41192 | 45918 | 50206 | 9951 |

ACEi – angiotensin-converter enzyme inhibitor, ARB - angiotensin II receptor blocker, BB - beta blocker, CCB - calcium channel blocker, MRA –mineralocorticoid receptor antagonists

The mean PDC for cardiovascular drug use in our cohort was 85.7% (median 93.0%) pre-pandemic and 87.0% (median 94.0%) during the pandemic and the proportion exhibiting good adherence (PDC>0.8) increased from 72.8% pre-pandemic to 75.4% during the pandemic (p<0.001). The PDC varied minimally across drug classes (eTable 2), but was higher for all drug classes during the pandemic compared to pre-pandemic (eTable 2). The proportions of patients exhibiting good adherence were higher during the pandemic than pre-pandemic for each of the drug classes we studied (Figure 2).

**Figure 2.**
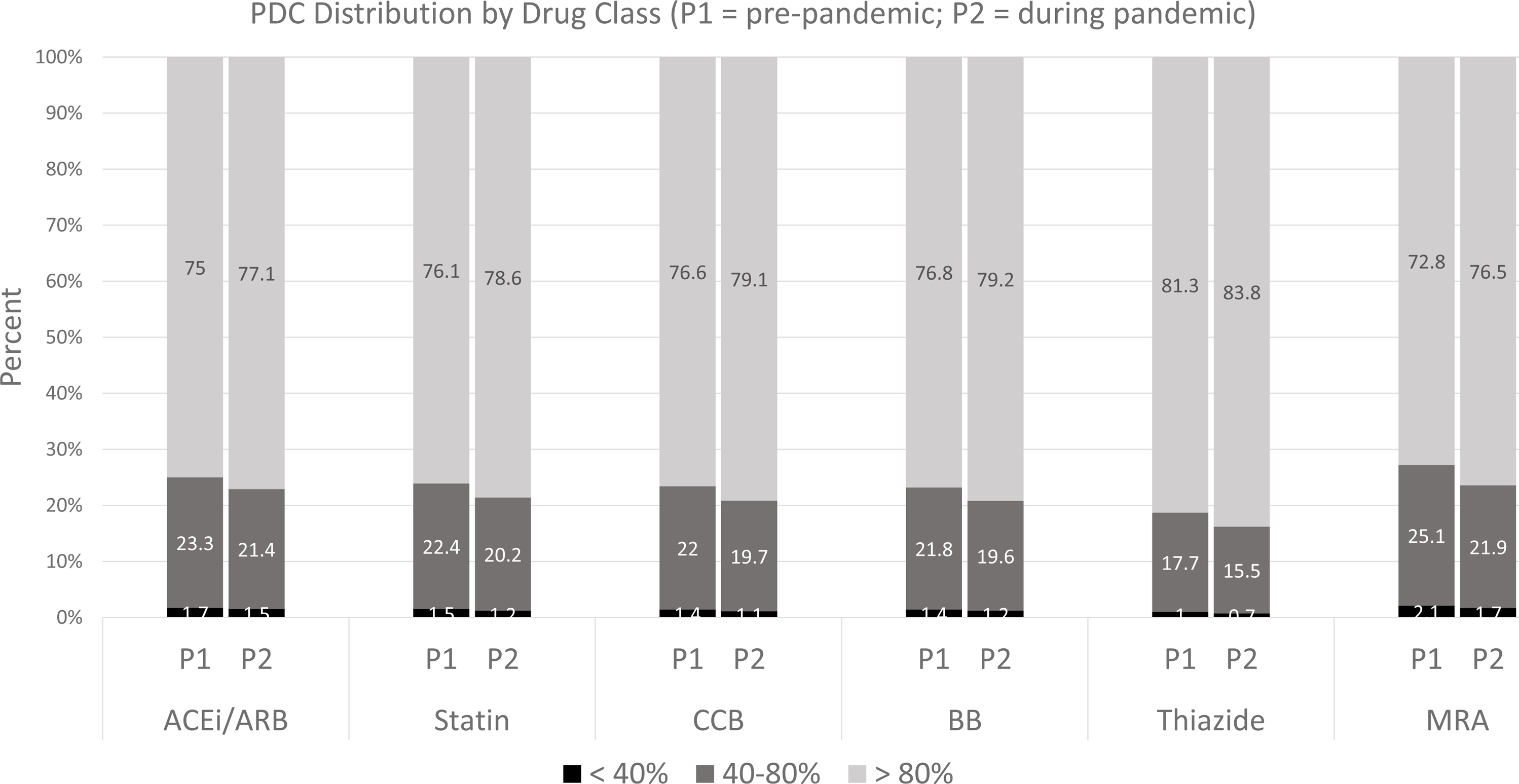
**PDC Distribution by Drug Class (P1 = pre-pandemic, P2 = pandemic)**

The multivariable logistic analyses with GEE revealed that the proportion of chronic users exhibiting “good adherence” (PDC>0.8) was significantly higher for all drug classes studied during the pandemic compared to prior (Table 3). During the pandemic, chronic users of cardiovascular drugs were 5% to 16% more likely to exhibit good adherence than they were in the pre-pandemic period (Table 3). Within each drug class-specific analysis, consistent predictors of better adherence were older age, higher socioeconomic status, rural residence, and a diagnosis of hypertension or diabetes mellitus. On the other hand, higher Charlson scores were associated with poorer adherence for all drug classes other than ACEi/ARB and the impact of sex and diagnoses of heart failure or ischemic heart disease varied between drug classes (Table 3).

**Table 3.**
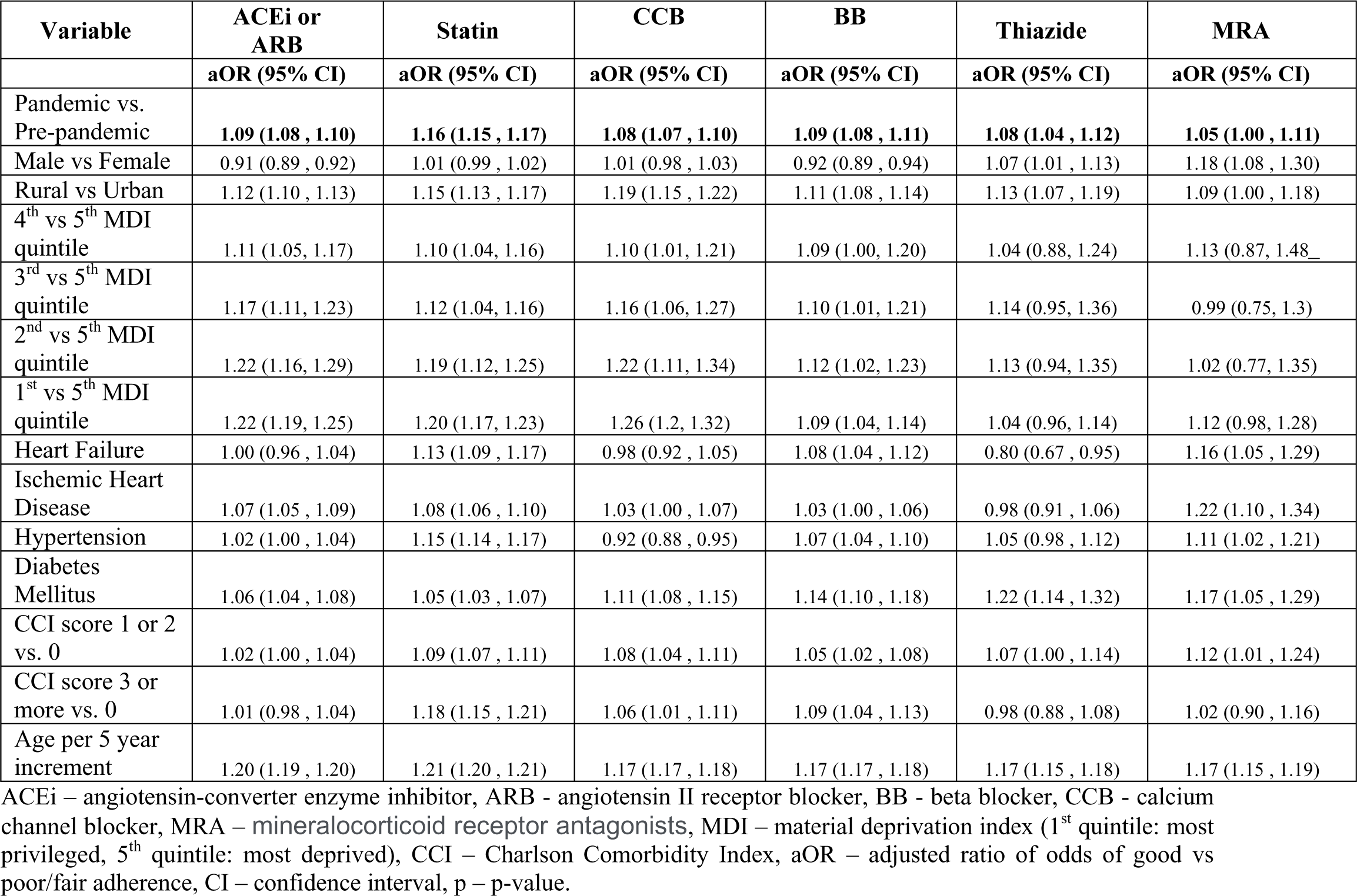
Predictors of good adherence (PDC ≥ 0.8) with each drug class.

## DISCUSSION

Our main finding is that although adherence patterns with cardiovascular drugs varied slightly between classes, adherence within each drug class for chronic users was higher during the COVID-19 pandemic compared to the two years prior. This is in contrast to earlier surveys[5–7] and a cohort study focusing on just the first 6 months of the pandemic[8] which suggested that the COVID-19 pandemic may have profound negative impacts on medication adherence.

A recent systematic review of 12 published studies of medication adherence during the pandemic found that most studies evaluated medications for respiratory diseases or inflammatory conditions, combined chronic, new, and transient users, and the data was not meta-analyzable given the variability in methods, metrics, and results.[12] For example, while one study showed improved adherence with inhaled bronchodilators for asthma patients during the pandemic,[13] another reported >20% drops in PDC for asthma control medications,[14] and a third found no change in refill rates for inhaled corticosteroids in patients with asthma during versus prior to the pandemic.[15]. However, the relevance of these studies to patients with cardiovascular diseases is questionable since there was an abundance of public health messaging emphasizing the dangers of COVID-19 in patients with pre-existing lung disease which may well have encouraged such patients to more rigorously adhere to their prescribed therapies. Two recently reported studies from Italy (only one of which was included in the systematic review mentioned above) reported minimal changes in adherence to statins, antihypertensive, or glucose lowering drugs during the first year of the pandemic, despite the fact Italy was particularly hard hit early in the pandemic.[16,17] A scoping review of barriers to antihypertensive medication adherence found that the COVID-19 pandemic did not introduce new barriers but did exacerbate pre-existing barriers of access and cost for disadvantaged populations.[18]

Although our study was able to capture all prescription medication dispensations from community pharmacies for all adults in the Canadian province of Alberta regardless of their disease or insurance status, there are some limitations to our work. For one, the databases analyzed included physician-assigned diagnoses but did not include details on clinical severity or physical measures like blood pressure or lipid levels, which precludes direct assessment of risk factor control or adequacy of disease management. Second, our estimates of adherence may be over-estimates since we excluded new users and individuals who did not fill at least 1 prescription per annum over all study years, and using pharmacy refills to monitor medication intake fails to capture primary non-adherence (the failure of a patient to fill even the first prescription).[19] Moreover, pharmacy refill data may not align with actual patient consumption and is unable to capture temporary stops, pill splitting, dosage adjustments using existing dispensed supplies, or the use of free samples. We may also have potentially under-captured comorbidities during the initial stages of COVID-19 when patient healthcare encounters decreased (although this is less than might be expected as we have previously documented that the frequency of outpatient encounters stayed largely the same pre/during pandemic because of the introduction of virtual billing codes allowing telephone follow-up).[20] Our study also does not assess changes in adherence associated with spikes and troughs in the number of COVID-19 cases, hospitalizations, or deaths which affected different areas at different times and may also have influenced adherence.

In conclusion, contrary to what surveys and very early studies suggested may happen, drug dispensations and patient adherence were not adversely affected by the COVID-19 pandemic, at least in the Canadian province of Alberta. In fact, we found that chronic users of cardiovascular drugs exhibited slightly higher adherence during the COVID-19 pandemic than prior to the pandemic. Thus, it would appear that processes initiated early in the pandemic to address the impact of societal lockdowns and reduced access to in-person outpatient care (principally the introduction of physician billing codes for the provision of virtual care) as well as the efforts of various professional societies, including the American Heart Association, to combat misinformation about the risks of cardiovascular drugs vis-à-vis COVID-19 had their intended effect and should help guide our responses to future pandemics.

## Data Availability

The data underlying this article was provided by the Government of Alberta under the terms of a research agreement and cannot be shared by the investigators under the Alberta Health Information Act. Inquiries regarding access to the data can be made to.

## Ethics Statement

This study was approved by the Health Research Ethics Board of the University of Alberta (Pro00115481) with a waiver of individual signed consent for this study.

## Acknowledgements

This study is based in part on data provided by Alberta Health and Alberta Health Services. We thank the Customer Relationship Management and Data Access Unit at Alberta Health for creating the linked database and the Alberta Strategy for Patient Oriented Research Unit for assistance with data analyses. The interpretation and conclusions are those of the researchers and do not represent the views of the Government of Alberta. Neither the Government of Alberta nor Alberta Health expressed any opinion in relation to this study.

## E-Appendix for

**eTable 1.** Case definitions for conditions and drug classes.

| **Condition** | **ICD-9** | **ICD-10-CA** |
| --- | --- | --- |
| Hypertension | 401 402 403 404 405 | I10 I11 I12 I13 I15 |
| Ischemic Heart Disease | 410 411 412 413 414 4292 | I20 I21 I22 I23 I24 I25 |
| Heart Failure | 3989 4020 4029 4040 4041 4049<br>4254 4255 4256 4257 4258 4259 428 | I099 I110 I130 I132 I255 I420 I425<br>I426 I427 I428 I429 I43 I50 P290 |
| Diabetes Mellitus | 250 | E10 E11 E12 E13 E14 |
| **Drug class** | **ATC** |  |
| ACEi or ARB | C09A C09B C09C C09D |  |
| Statin | C10AA C10BA C10BX |  |
| CCB | C08 |  |
| BB | C07 |  |
| Thiazide | C03AA C03AB |  |
| MRA | C03DA |  |

ICD - International Classification of Diseases, -9 – 9<sup>th</sup> revision, -10-CA – 10<sup>th</sup> revision clinical modification for Canada, ATC – anatomical therapeutic chemical, ACEi – angiotensin-converter enzyme inhibitor, ARB - angiotensin II receptor blocker, BB - beta blocker, CCB - calcium channel blocker, MRA – mineralocorticoid receptor antagonists

**eTable 2.** Characteristics of chronic users of each cardiovascular drug class (columns are not mutually exclusive)

| Factor | Pre-pandemic [Mar 15 2018 - Mar 14 2020] |  |  |  |  |  | Pandemic [Mar 15 2020 - Mar 14 2022] |  |  |  |  |  |
| --- | --- | --- | --- | --- | --- | --- | --- | --- | --- | --- | --- | --- |
|  | ACEi or ARB | Statin | CCB | BB | Thiazide | MRA | ACEi or ARB | Statin | CCB | BB | Thiazide | MRA |
| **N** | **370945** | **295421** | **113551** | **115011** | **29465** | **9951** | **370945** | **295421** | **113551** | **115011** | **29465** | **9951** |
| **Age (mean, SD)** | 62.3 (12.1) | 63.7 (10.9) | 64.7 (12.5) | 65.5 (12.5) | 64.0 (11.5) | 59.9 (15.2) | 64.3 (12.1) | 65.7 (10.9) | 66.7 (12.5) | 67.5 (12.5) | 66.0 (11.5) | 61.9 (15.2) |
| **Age 65+** | 162050 (43.7) | 141336 (47.8) | 59547 (52.4) | 64345 (55.9) | 14503 (49.2) | 4139 (41.6) | 185973 (50.1) | 162611 (55.0) | 66479 (58.5) | 71481 (62.2) | 16506 (56.0) | 4650 (46.7) |
| **Female** | 170420 (45.9) | 119573 (40.5) | 54516 (48.0) | 49369 (42.9) | 17717 (60.1) | 5505 (55.3) | 170420 (45.9) | 119573 (40.5) | 54516 (48.0) | 49369 (42.9) | 17717 (60.1) | 5505 (55.3) |
| **Urban** | 221028 (59.6) | 181270 (61.4) | 69413 (61.1) | 67097 (58.3) | 15954 (54.1) | 5595 (56.2) | 220405 (59.4) | 180681 (61.2) | 69259 (61.0) | 67031 (58.3) | 15904 (54.0) | 5603 (56.3) |
| **Material Deprivation Index 5<sup>th</sup> quintile (most deprived)** | 83187 (22.4) | 64755 (21.9) | 26229 (23.1) | 26248 (22.8) | 6312 (21.4) | 2154 (21.6) | 81697 (22.0) | 63644 (21.5) | 25769 (22.7) | 25874 (22.5) | 6193 (21.0) | 2121 (21.3) |
| **4<sup>th</sup> quintile** | 81260 (21.9) | 63574 (21.5) | 24807 (21.8) | 25277 (22.0) | 6509 (22.1) | 2253 (22.6) | 80728 (21.8) | 63110 (21.4) | 24648 (21.7) | 25032 (21.8) | 6480 (22.0) | 2214 (22.2) |
| **3<sup>rd</sup> quintile** | 69232 (18.7) | 54817 (18.6) | 20753 (18.3) | 21020 (18.3) | 5566 (18.9) | 1817 (18.3) | 69308 (18.7) | 54778 (18.5) | 20811 (18.3) | 21049 (18.3) | 5571 (18.9) | 1814 (18.2) |
| **2<sup>nd</sup> quintile** | 61665 (16.6) | 49828 (16.9) | 18508 (16.3) | 18490 (16.1) | 4851 (16.5) | 1614 (16.2) | 62414 (16.8) | 50512 (17.1) | 18655 (16.4) | 18614 (16.2) | 4911 (16.7) | 1596 (16.0) |
| **1<sup>st</sup> quintile (most privileged)** | 57042 (15.4) | 47941 (16.2) | 17066 (15.0) | 17132 (14.9) | 4705 (16.0) | 1523 (15.3) | 57594 (15.5) | 48322 (16.4) | 17229 (15.2) | 17263 (15.0) | 4707 (16.0) | 1560 (15.7) |
| **Missing** | 18559 (5.0) | 14506 (4.9) | 6188 (5.4) | 6844 (6.0) | 1522 (5.2) | 590 (5.9) | 19204 (5.2) | 15055 (5.1) | 6439 (5.7) | 7179 (6.2) | 1603 (5.4) | 646 (6.5) |
| **CCI Score Mean (SD)** | 0.9 (1.4) | 1.0 (1.5) | 1.0 (1.5) | 1.3 (1.8) | 0.7 (1.2) | 1.8 (2.2) | 1.0 (1.5) | 1.1 (1.6) | 1.1 (1.6) | 1.4 (1.9) | 0.8 (1.2) | 1.8 (2.2) |
| **CCI Score group** |  |  |  |  |  |  |  |  |  |  |  |  |
| **0** | 209852 (56.6) | 145575 (49.3) | 62356 (54.9) | 54967 (47.8) | 18732 (63.6) | 3954 (39.7) | 195769 (52.8) | 136932 (46.4) | 57838 (50.9) | 52277 (45.5) | 17801 (60.4) | 3935 (39.5) |
| **1, 2** | 121608 (32.8) | 111266 (37.7) | 36113 (31.8) | 37914 (33.0) | 8456 (28.7) | 3073 (30.9) | 126931 (34.2) | 113223 (38.3) | 37755 (33.2) | 38288 (33.3) | 9009 (30.6) | 3127 (31.4) |
| **3+** | 39485 (10.6) | 38580 (13.1) | 15082 (13.3) | 22130 (19.2) | 2277 (7.7) | 2924 (29.4) | 48245 (13.0) | 45266 (15.3) | 17958 (15.8) | 24446 (21.3) | 2655 (9.0) | 2889 (29.0) |
| **Comorbidities based on all encounters in the 24 months up to time of prescription:** |  |  |  |  |  |  |  |  |  |  |  |  |
| **Heart Failure (n,%)** | 15847 (4.3) | 13753 (4.7) | 4471 (3.9) | 15122 (13.1) | 622 (2.1) | 3705 (37.2) | 18841 (5.1) | 16415 (5.6) | 5320 (4.7) | 16798 (14.6) | 631 (2.1) | 3550 (35.7) |
| **Ischemic Heart Disease (n,%)** | 64613 (17.4) | 69114 (23.4) | 18898 (16.6) | 48185 (41.9) | 3372 (11.4) | 3194 (32.1) | 67119 (18.1) | 69987 (23.7) | 19225 (16.9) | 47062 (40.9) | 3475 (11.8) | 3029 (30.4) |
| **Hypertension (n,%)** | 321680 (86.7) | 194618 (65.9) | 102047 (89.9) | 87834 (76.4) | 25502 (86.6) | 6312 (63.4) | 326215 (87.9) | 202818 (68.7) | 102038 (89.9) | 89186 (77.5) | 25433 (86.3) | 6196 (62.3) |
| **Diabetes Mellitus (n,%)** | 119026 (32.1) | 113711 (38.5) | 36849 (32.5) | 37397 (32.5) | 7512 (25.5) | 3058 (30.7) | 129269 (34.8) | 121317 (41.1) | 39896 (35.1) | 40415 (35.1) | 8357 (28.4) | 3336 (33.5) |
| **Drug Class (n,%):** |  |  |  |  |  |  |  |  |  |  |  |  |
| **ACEi or ARB** | 370945 (100.0) | 202484 (68.5) | 84981 (74.8) | 84163 (73.2) | 20930 (71.0) | 6524 (65.6) | 370945 (100.0) | 205862 (69.7) | 84968 (74.8) | 83796 (72.9) | 21152 (71.8) | 6388 (64.2) |
| **Statin** | 210038 (56.6) | 295421 (100.0) | 63795 (56.2) | 78018 (67.8) | 14766 (50.1) | 5019 (50.4) | 223275 (60.2) | 295421 (100.0) | 67362 (59.3) | 79763 (69.4) | 15668 (53.2) | 5174 (52.0) |
| **CCB** | 115366 (31.1) | 79238 (26.8) | 113551 (100.0) | 35643 (31.0) | 10047 (34.1) | 2458 (24.7) | 124235 (33.5) | 84506 (28.6) | 113551 (100.0) | 36430 (31.7) | 10480 (35.6) | 2267 (22.8) |
| **BB** | 93938 (25.3) | 87054 (29.5) | 30390 (26.8) | 115011 (100.0) | 6720 (22.8) | 5474 (55.0) | 99143 (26.7) | 89772 (30.4) | 31616 (27.8) | 115011 (100.0) | 6941 (23.6) | 5446 (54.7) |
| **Thiazide** | 51203 (13.8) | 31219 (10.6) | 18943 (16.7) | 13884 (12.1) | 29465 (100.0) | 590 (5.9) | 47308 (12.8) | 28274 (9.6) | 17078 (15.0) | 12321 (10.7) | 29465 (100.0) | 423 (4.3) |
| **MRA** | 12483 (3.4) | 9639 (3.3) | 3886 (3.4) | 9736 (8.5) | 365 (1.2) | 9951 (100.0) | 15699 (4.2) | 11909 (4.0) | 4812 (4.2) | 11314 (9.8) | 435 (1.5) | 9951 (100.0) |
| **Adherence:** |  |  |  |  |  |  |  |  |  |  |  |  |
| **Mean PDC (SD)** | 90.4 (14.0) | 88.9 (15.1) | 90.4 (14.2) | 90.8 (14.2) | 90.4 (14.1) | 88.0 (16.4) | 91.4 (13.4) | 90.4 (14.1) | 91.3 (13.7) | 91.7 (13.6) | 91.4 (13.2) | 88.8 (15.8) |
| **Median PDC (IQR)** | 97 (87, 100) | 95 (85, 99) | 97 (87, 100) | 97 (88, 100) | 97 (87, 99) | 95 (83, 99) | 97 (89, 100) | 96 (88, 99) | 97 (89, 100) | 98 (90, 100) | 97 (89, 100) | 96 (84, 99) |
| **PDC < 0.4 (poor)** | 6286 (1.7) | 4225 (1.4) | 1572 (1.4) | 1527 (1.3) | 291 (1.0) | 207 (2.1) | 5446 (1.5) | 3688 (1.2) | 1304 (1.1) | 1333 (1.2) | 217 (0.7) | 166 (1.7) |
| **PDC 0.4 to 0.8 (fair)** | 86634 (23.4) | 66315 (22.4) | 24975 (22.0) | 25120 (21.8) | 5221 (17.7) | 2497 (25.1) | 79560 (21.4) | 59573 (20.2) | 22377 (19.7) | 22566 (19.6) | 4556 (15.5) | 2175 (21.9) |
| **PDC ≥ 0.8 (good)** | 278025 (75.0) | 224881 (76.1) | 87004 (76.6) | 88364 (76.8) | 23953 (81.3) | 7247 (72.8) | 285939 (77.1) | 232160 (78.6) | 89870 (79.1) | 91112 (79.2) | 24692 (83.8) | 7610 (76.5) |

ACEi – angiotensin-converter enzyme inhibitor, ARB - angiotensin II receptor blocker, BB - beta blocker, CCB - calcium channel blocker, MRA – mineralocorticoid receptor antagonists, PDC – proportion of days covered, SD – standard deviation, IQR – interquartile range, CCI – Charlson Comorbidity Index.

## REFERENCES

1. Chowdhury R, Khan H, Heydon E, et al. Adherence to cardiovascular therapy: a meta-analysis of prevalence and clinical consequences. Eur Heart J. 2013 Oct;34(38):2940–8.

2. Simpson SH, Eurich DT, Majumdar SR, Padwal RS, Tsuyuki RT, Varney J, Johnson JA. A meta-analysis of the association between adherence to drug therapy and mortality. BMJ. 2006;333:15.

3. Cutler RL, Fernandez-Llimos F, Frommer M, Benrimoj C, Garcia-Cardenas V. Economic impact of medication non-adherence by disease groups: a systematic review. BMJ Open. 2018;8(1):e016982. doi: 10.1136/bmjopen-2017-016982.

4. Roebuck MC, Kaestner RJ, Dougherty JS. Impact of Medication Adherence on Health Services Utilization in Medicaid. Med Care.2018;56(3):266–273.

5. Chagué F, Boulin M, Eicher JC, Bichat F, Saint Jalmes M, Cransac-Miet A, Soudry-Faure A, Danchin N, Cottin Y, Zeller M. Impact of lockdown on patients with congestive heart failure during the coronavirus disease 2019 pandemic. ESC Heart Fail. 2020 Sep 30;7(6):4420–3

6. Santi RL, Márquez MF, Piskorz D, Saldarriaga C, Lorenzatti A, Wyss F, Martín AV, Perales JS, Arcela JC, de Lourdes Rojas Gimon E, Sambadaro G, Perez GE, Mendoza I, Lanas F, Flores R, Liprandi AS, Alexander B, Baranchuk A. Ambulatory Patients with Cardiometabolic Disease and Without Evidence of COVID-19 During the Pandemic. The CorCOVID LATAM Study. Glob Heart. 2021 Feb 17;16(1):15

7. Ismail H, Marshall VD, Patel M, Tariq M, Mohammad RA. The impact of the COVID-19 pandemic on medical conditions and medication adherence in people with chronic diseases. J Am Pharm Assoc (2003). 2021 Nov 15:S1544-3191(21)00474-X. doi: 10.1016/j.japh.2021.11.013. Epub ahead of print. PMID: 34844885; PMCID: PMC8591859.

8. Egan BM, Sutherland SE, Macri CI, et al. Association of Baseline Adherence to Antihypertensive Medications With Adherence After Shelter-in-Place Guidance for COVID-19 Among US Adults. JAMA Netw Open. 2022;5(12):e2247787.

9. Halpern MT, Khan ZM, Schmier JK, Burnier M, Caro JJ, Cramer J, et al. Recommendations for evaluating compliance and persistence with hypertension therapy using retrospective data. Hypertension 2006;47(6):1039–48.

10. Tonelli M, Wiebe N, Fortin M, et al. for the Alberta Kidney Disease Network. Methods for identifying 30 chronic conditions: application to administrative data. BMC Medical Informatics and Decision Making 2015;15:31.

11. Schneeweiss S, Wang PS, Avorn J, Glynn RJ. Improved comorbidity adjustment for predicting mortality in Medicare populations. Health Serv Res. 2003 Aug;38(4):1103–20.

12. Olmastroni E, Galimberti F, Tragni E, Catapano AL, Casula M. Impact of COVID-19 Pandemic on Adherence to Chronic Therapies: A Systematic Review. Int J Environ Res Public Health. 2023 Feb 21;20(5):3825.

13. Kaye L, Theye B, Smeenk I, Gondalia R, Barrett MA, Stempel DA. Changes in medication adherence among patients with asthma and COPD during the COVID-19 pandemic. J Allergy Clin Immunol Pract 2020;8:2384–2385.

14. Ramey O.L., Silva Almodovar A., Nahata M.C. Medication adherence in Medicare-enrolled older adults with asthma before and during the coronavirus disease 2019 pandemic. Ann. Allergy Asthma Immunol. 2022;128:561–567

15. Rodríguez I, López-Caro JC, Gonzalez-Carranza S, Cerrato ME, De Prado MM, Gomez-Molleda F, Pinel M, Saiz MT, Fuentes C, Barreiro E, Santibáñez M. Adherence to inhaled corticosteroids in patients with asthma prior to and during the COVID-19 pandemic. Sci Rep. 2023 Aug 11;13(1):13086

16. Romagnoli A., Santoleri F., Costantini A. The impact of COVID-19 on chronic therapies: The Pescara (ASL) local health authority experience in Italy. Curr. Med. Res. Opin. 2022;38:311–316.

17. Casula M, Galimberti F, Iommi M, Olmastroni E, Rosa S, Altini M, Catapano AL, Tragni E, Poluzzi E. Impact of the COVID-19 Pandemic on the Therapeutic Continuity among Outpatients with Chronic Cardiovascular Therapies. Int J Environ Res Public Health. 2022 Sep 24;19(19):12101.

18. Foster M, Etchin A, Pope C, Hartmann CW, Emidio O, Bosworth HB. The Impact of COVID-19 on Hypertension and Hypertension Medication Adherence Among Underrepresented Racial and Ethnic Groups: A Scoping Review. Curr Hypertens Rep. 2023 Nov;25(11):385–394.

19. Zeitouny S, Cheng L, Wong ST, Tadrous M, McGrail K, Law MR. Prevalence and predictors of primary nonadherence to medications prescribed in primary care. CMAJ. 2023 Aug 8;195(30):E1000–E1009.

20. McAlister FA, Hsu Z, Dong Y, Tsuyuki RT, van Walraven C, Bakal J. Frequency and type of outpatient visits for patients with cardiovascular ambulatory care sensitive conditions during the COVID-19 pandemic and subsequent outcomes: A retrospective cohort study. JAHA 2023;12:e027922.

